# COVID-19 disease progression according to initial symptoms. A telemedicine cohort study

**DOI:** 10.1101/2022.01.03.22268669

**Authors:** Karla Murillo-Villanueva, Blanca Velázquez-Hernández, José A. Jácome-Mondragón, Judit J. Cervantes-Llamas, Juan O. Talavera

## Abstract

**Objective:** COVID-19 progression to severe or critical illness may be related to initial clinical presentation. Main objective was to identify initial symptoms related to highest risk of disease progression, in mild or moderate suspected or confirmed COVID-19 patients or in asymptomatic subjects in contact with a recently diagnosed patient.

**Design and methods:** Historic cohort study of Mexican patients with suspected or confirmed mild or moderate COVID-19 or asymptomatic subjects in recent contact with positive patients. They sought medical attention in “Centro Médico ABC” or claimed for remote attention, and daily telemedicine follow up until recovery or illness progression, from April 17th to October 08th 2020. Data excerpted for analysis were sex, age, body mass index, comorbidities, and signs, and symptoms presented in first day of disease manifestations and during follow up. We used logistic regression to identify initial symptoms associated with progression disease and through a conjunctive consolidation analysis a symptom index was created.

**Results:** 120 of 1635 patients (7.2%) had clinical progression disease. By logistic regression we found as initial symptoms related to progression: fever OR 3 (1.89-4.77, p<0.001), cough OR 2.34 (1.56-3.52, p<0.001), myalgias or arthralgias OR 1.69 (1.09-2.63, p=0.018), and fatigue OR 1.65 (1.08-2.53, p=0.019). Conjunctive consolidation was processed with the previous symptoms, and a 3 groups score resulted C-19PAIS Index: 1) Fever with cough or fever with fatigue, with a probability of progression disease of 29% (31/106 patients), 2) Fever or cough or fatigue or cough with fatigue, 10.7% (66/615 patients) and 3) No fever, no cough, no fatigue, 2% (23/914).

**Conclusions:** Initial symptoms predict clinical progression in COVID-19 patients.

## Introduction

The new SARS CoV-2 (severe acute respiratory syndrome coronavirus 2), following the onset in 2019 (1) of multiple cases of pneumonia of unknown etiology in Hubei Province, China, originated the coronavirus disease-2019 (COVID-19) pandemic(2).

COVID-19 may present as asymptomatic, mild, moderate, severe, or critical disease, with higher mortality in severe and critical cases. Most patients with COVID-19 initiate as mild or moderate disease(3-5), among them between 3% to 20% (5-7) progress to severe and critical disease.

Risk factors for developing severe or critical disease are male gender, older, presenting comorbidities, being overweight or obese. Also, presenting abnormal radiological manifestations and a short or long period between onset and progression of symptoms is associated with a worst prognosis (3, 7-9).

The pandemic provoked a change in current medicine, increasing the demand for telemedicine to avoid exposure in hospitals to COVID-19 patients. It included general disease patients, as well as COVID-19 suspected mild or moderate disease patients or subjects in recent contact with positive COVID-19 patients (10). In view of this situation, it is of the most importance to identify which diseased patients or individuals under exposure may present the highest risk of clinical disease progression.

The aim of this study is to determine which onset symptoms among mild and moderated diagnosed or suspected COVID-19 patients or subjects in recent contact with positive COVID-19 patients represent the highest risk of disease progression.

## Materials and methods

### Study design and data source

This is a historic cohort study using data collected from a telemedicine program available to Mexican patients in response to the emerging pandemic. This program provides remote assessment by general practitioners and specialists to patients at home. Patients with confirmed or suspected COVID-19 diagnosis or asymptomatic in recent contact with positive COVID-19 patients, who sought medical attention in “Centro Médico ABC (private and community health service)”, or claimed for remote attention, which were send or kept at home for daily telemedicine follow up until recovery for a media of 14 days or illness progression; in such cases patients were referred to hospital evaluation.

Initial evaluation at hospital or self-assessed data at home, such as initial symptoms or signs, date of onset, SpO_2_ (when available) and body temperature (when available) and follow up data obtained by interview were recorded in the telemedicine program records of COVID-19 patients of the Centro Medico American British Cowdray, in Mexico City. The data in this article are from April 17^th^ 2020 to October 08^th^ 2020. The Institutional Review Board of CMABC approved this study (Protocol approval ABC-20-93).

### Variable definition

We included suspected clinical COVID-19 subjects as it was determined by government health ministry’s operational definition, as a person presenting two of the following manifestations: cough, fever or headache accompanied by at least one of dyspnea, arthralgias, myalgias, odynophagia, rhinorrhea, conjunctivitis, or chest pain. This definition changed during the study period, as presenting at least one of these: headache, fever, cough, or dyspnea adding any of myalgia, arthralgia, odynophagia, chills, chest pain, rhinorrhea, anosmia, dysgeusia or conjunctivitis. Also were included any asymptomatic person with recent contact with COVID-19 suspected or positive patients. All of them with an age >18 years, not pregnant, with an oxygen saturation measurement ≥90% when evaluated at hospital, and in those patients at home that referred dyspnea an evaluation of oxygen saturation measurement ≥90% was necessary.

Given that the government exhorted patients to self-isolate unless they presented serious symptoms (dyspnea or persistent fever), many of them did not look for hospital attention and chose to stay at home, consequently their follow-up took place at their home and did not have a PCR study.

Initial clinical symptoms and signs evaluated according to the first day of presentation, referred by patients were: headache, fever, cough, dyspnea, myalgia or arthralgia, odynophagia, fatigue or general malaise, rhinorrhea, anosmia or dysgeusia and diarrhea, and measured at hospital or home when available: body temperature and SpO_2_%.

Other evaluated parameters were demographic characteristics such as age, gender, body weight and height (referred by the patient), comorbidities like diabetes, hypertension, chronic obstructive pulmonary disease, asthma, chronic kidney diseases, tobacco use, immunocompromised. RT-PCR testing for SARS-CoV-2 reported as positive, negative, or not tested.

Patients that during the follow up referred increase in dyspnea or prolonged fever, general malaise, or cough, were send to medical evaluation at hospital, and disease progression diagnoses was stablished by meeting the two following criteria: a) oxygen saturation less than 90% in room air(11), and b) shortness of breath or pneumonia (12).

### Data analysis: Coding and substitution of variables

Age was coded in two groups, > or ≤ 60 years old; weight and height were converted into body mass index (BMI), coded as healthy <25 kg/m^2^, overweight as 25.0-29.9 kg/m^2^, and obese ≥ 30 kg/m^2^. Temperature was registered in °C and coded into two groups, ≥ or <38°C, SpO_2_ in % by pulsioximeter (self-reported when available or data recollected from medical evaluation) and coded as ≥ or < 94%. Other signs and all symptoms, and comorbidities were coded as present or absent. Finally, PCR SARS CoV-2 test result was coded as positive, negative, or not tested.

Time of progression was the period of days among onset of initial symptoms and the presence of aggravation.

Missing data was substituted using the median for age (34 patients, 2.1%). And a missing stratum was included for the following variables just to keep them in the multivariable analysis, body mass index (missing 702, 42.9%), temperature (missing 618, 37.8%), and SpO_2_ (missing 729, 44.6%).

### Sample size calculation

The sample size calculation was based on the requirements for a multivariable model (events per variable, EPV); considering that 6.2% was estimated to present the outcome of clinical progression (13), and following the assumption of 10-20 events per variable (14), and that the study has 8 parameters (symptoms), we calculated a minimum required sample size of 1324 patients, expecting 80 patients with progressive illness; and a maximum of 2648.

### Statistical analysis

Demographic characteristics, comorbid conditions, initial symptoms, and signs were compared against outcome (progression VS no progression), in a bivariate analysis by a Chi-square test.

Then a multivariable model contrasting significant initial symptoms against outcome was carried out in 5 steps in order two adjust for different group variables: step1) Only symptoms, step2) step 1 variables plus age and gender, step3) step2 variables plus comorbidities (overweight, obesity, hypertension, diabetes, and tobacco use). In step4 SpO_2_ was added and then int step5 PCR testing result.

Each logistic regression model is presented with its Odds Ratio (OR), 95% Confidence Interval (CI95%), and its “p value”.

Finally, those symptoms that maintained statistical significance trough multivariate analysis were consolidated (15, 16), and a Progression According to Initial Symptom Index (C-19PAIS Index) was created, its different stratums were contrasted against age and comorbidities.

Statistical significance was set at p<0.05 and performed with SPSS version 27.0

## Results

The study included 1635 patients. Women were 60.1%, median age of 36 years (percentile (pc) 25-75 was 29-47), 91.4% were younger than 60 years old. Of the 933 that registered body weight and height, the median BMI was 26.1 kg/m^2^ (pc25-75, 23.5-29.7), 39.8% were healthy, 36.9% with overweight and 23.4 obese. The most common comorbidity was hypertension in 9.7%, followed by tobacco use in 7.5%, and diabetes mellitus in 5%.

The most frequent initial symptom was headache in 42.3%, followed by myalgia or arthralgias (33.6%), sore throat (33.3%), fatigue (25.7%), cough (22.3%), rhinorrhea (17.1%), diarrhea (12.7%), anosmia or dysgeusia (11.6%), fever (10.6%), dyspnea or chest pain (7.6%). The initial temperature recoded was available in 1017 patients with a median of 36.5° C (pc15-75, 36.0-37.5°C), ≤ 37.4°C in 74%, between 37.5-37.9°C in 8.8%, and ≥38.0°C in 17.2%. Patients with initial oxygen saturation were 802, with a median SpO2 of 95% (pc25-75, 94-96%), of this, 77.3% had ≥94%; 906 had a PCR for SARS CoV 2 test available; 54.7% positive result. Characteristics of all patients are summarized in Table 1.

**Table 1.** Patient demographic characteristics and initial symptoms and signs.

| Variable | N = 1635, n (%) |
| --- | --- |
| Female | 983 (60.1) |
| Male | 652 (39.9) |
| Age<sup>a</sup> | 36 (29-47) |
| <60 | 1494 (91.4) |
| ≥60 | 141 (8.6) |
| BMI<sup>a</sup> | 26.1 (23.5-29.7) |
| <25 kg/m<sup>2</sup> | 371 (39.8) |
| 25-29.9 kg/m<sup>2</sup> | 344 (36.9) |
| ≥30 kg/m<sup>2</sup> | 218 (23.4) |
| **Comorbidities** |  |
| Hypertension | 158 (9.7) |
| Tobacco Use | 123 (7.5) |
| Diabetes Mellitus | 81 (5) |
| Asthma | 37 (2.3) |
| Immunosuppression | 16 (1) |
| COPD | 14 (0.9) |
| CKD | 5 (0.3) |
| **Initial symptoms** |  |
| Headache | 692 (42.3) |
| Myalgias / Arthralgias | 549 (33.6) |
| Odynophagia | 544 (33.3) |
| Fatigue / General malaise | 420 (25.7) |
| Cough | 365 (22.3) |
| Rhinorrhea | 279 (17.1) |
| Diarrhea | 208 (12.7) |
| Anosmia / Ageusia | 190 (11.6) |
| Fever | 174 (10.6) |
| Dyspnea | 128 (7.8) |
| **Initial Signs** |  |
| Body Temperature °C<sup>a</sup> | 36.5 (36– 37.5) |
| ≤37.4 | 753 (74) |
| 37,5-37,9 | 90 (8.8) |
| ≥38 | 174 (17.2) |
| SpO2 %<sup>a</sup> | 95 (94-96) |
| ≥94 | 620 (77.3) |
| ≤93 | 182 (22.7) |
| PCR test result |  |
| Negative | 496 (54.7) |
| Positive | 410 (45.3) |

<sup>a</sup> Data is represented in median and Interquartile Range (IQR). COPD = chronic obstructive pulmonary disease, CKD = chronic kidney disease.

Table 2A shows the differences of demographic and comorbidity of patients with progression against (vs) No-progression (stable) disease. Progression disease occurred in 120 patients (7.3%), the median time was 7 days (pct 25-75, 5-9). Progression disease occurs in 9.4% male vs 6% of women, with a median age 44 (34-54.5) vs 36 (28-46) in no-progression disease. Also, among progression disease BMI was <25 in 5.1%, 25-29.9 in 8.7% and ≥30 in14.7%, and the presence vs absent were for: hypertension 13.3% vs 6.7%, diabetes mellitus 17.3% vs 6.8%, immunosuppression 31.1% vs 7.1%, and chronical kidney disease 60% vs 7.2%. All of them with a p value <0.05.

**Table 2.A.**
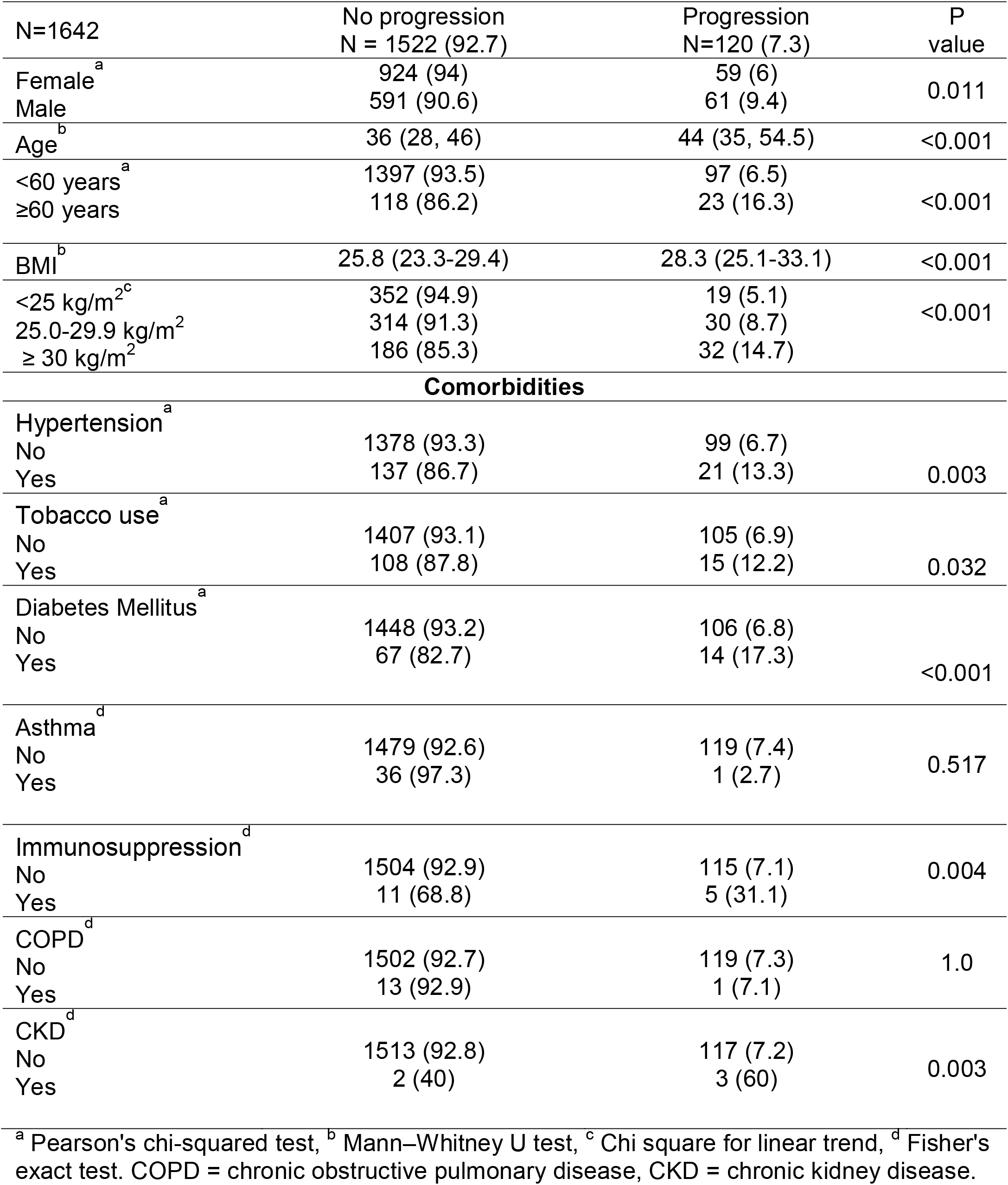
Patient demographic characteristics according to disease progression.

| N=1642 | No progression<br>N = 1522 (92.7) | Progression<br>N=120 (7.3) | P<br>value |
| --- | --- | --- | --- |
| Female<sup>a</sup> | 924 (94) | 59 (6) | 0.011 |
| Male | 591 (90.6) | 61 (9.4) |  |
| Age<sup>b</sup> | 36 (28, 46) | 44 (35, 54.5) | <0.001 |
| <60 years<sup>a</sup> | 1397 (93.5) | 97 (6.5) | <0.001 |
| ≥60 years | 118 (86.2) | 23 (16.3) |  |
| BMI<sup>b</sup> | 25.8 (23.3-29.4) | 28.3 (25.1-33.1) | <0.001 |
| <25 kg/m<sup>2c</sup> | 352 (94.9) | 19 (5.1) | <0.001 |
| 25.0-29.9 kg/m<sup>2</sup> | 314 (91.3) | 30 (8.7) |  |
| ≥ 30 kg/m<sup>2</sup> | 186 (85.3) | 32 (14.7) |  |
| Comorbidities |  |  |  |
| Hypertension<sup>a</sup> |  |  | 0.003 |
| No | 1378 (93.3) | 99 (6.7) |  |
| Yes | 137 (86.7) | 21 (13.3) |  |
| Tobacco use<sup>a</sup> |  |  | 0.032 |
| No | 1407 (93.1) | 105 (6.9) |  |
| Yes | 108 (87.8) | 15 (12.2) |  |
| Diabetes Mellitus<sup>a</sup> |  |  | <0.001 |
| No | 1448 (93.2) | 106 (6.8) |  |
| Yes | 67 (82.7) | 14 (17.3) |  |
| Asthma<sup>d</sup> |  |  | 0.517 |
| No | 1479 (92.6) | 119 (7.4) |  |
| Yes | 36 (97.3) | 1 (2.7) |  |
| Immunosuppression<sup>d</sup> |  |  | 0.004 |
| No | 1504 (92.9) | 115 (7.1) |  |
| Yes | 11 (68.8) | 5 (31.1) |  |
| COPD<sup>d</sup> |  |  | 1.0 |
| No | 1502 (92.7) | 119 (7.3) |  |
| Yes | 13 (92.9) | 1 (7.1) |  |
| CKD<sup>d</sup> |  |  | 0.003 |
| No | 1513 (92.8) | 117 (7.2) |  |
| Yes | 2 (40) | 3 (60) |  |

<sup>a</sup> Pearson's chi-squared test, <sup>b</sup> Mann–Whitney U test, <sup>c</sup> Chi square for linear trend, <sup>d</sup> Fisher's exact test. COPD = chronic obstructive pulmonary disease, CKD = chronic kidney disease.

Table 2B shows initial symptoms association with progression disease. The most relevant symptoms associated with progression disease according with its difference between present vs absent were: 1) fever with a gap of 16.2% (21.8 vs 5.6%), followed by 2) cough 9.6% (14.8% vs 5.2%), 3) fatigue/general malaise 7.5% (12.9 vs 5.4%), 4) myalgias/arthralgias 7.3 (12.2% vs 4.9%), and 5) headache 4.1 (5.6% vs 9.7%). And among signs: temperature ≥38.0°C in 21.8%, between 37.5-37.9 in 16.7% and ≤37.4 in 7%, SpO2 ≤93% in 23.1%, and ≥94% in 7.6%. All of them with a p≤0.001.

**Table 2.B.** Initial symptoms and signs according to disease progression.

| Variable<br>N=1642 | No progression<br>N = 1522 (92.7) | Progression<br>N=120 (7.3) | P<br>value |
| --- | --- | --- | --- |
| Initial Symptoms |  |  |  |
| Headache<sup>a</sup> |  |  |  |
| No | 890 (94.4) | 53 (5.6) | 0.002 |
| Yes | 625 (90.3) | 67(9.7) |  |
| Myalgias / Arthralgias<sup>a</sup> |  |  |  |
| No | 1033 (95.1) | 53 (4.9) | <0.001 |
| Yes | 482 (87.8) | 67 (12.2) |  |
| Odynophagia<sup>a</sup> |  |  |  |
| No | 1016 (93.1) | 75 (6.9) | 0.307 |
| Yes | 499 (91.7) | 45 (8.3) |  |
| Fatigue / General Malaise<sup>a</sup> |  |  |  |
| No | 1149 (94.6) | 66 (5.4) | <0.001 |
| Yes | 366 (87.1) | 54 (12.9) |  |
| Cough<sup>a</sup> |  |  |  |
| No | 1204 (94.8) | 66 (5.2) | <0.001 |
| Yes | 311 (85.2) | 54 (14.8) |  |
| Rhinorrhea<sup>a</sup> |  |  |  |
| No | 1260 (92.9) | 96 (7.1) | 0.374 |
| Yes | 255 (91.4) | 24 (8.6) |  |
| Diarrhea<sup>a</sup> |  |  |  |
| No | 122 (92.6) | 105 (7.4) | 0.94 |
| Yes | 193 (92.8) | 15 (7.2) |  |
| Anosmia / Ageusia<sup>a</sup> |  |  |  |
| No | 1341 (92.8) | 104 (7.2) | 0.543 |
| Yes | 174 (91.6) | 16 (8.4) |  |
| Fever<sup>a</sup> |  |  |  |
| No | 1379 (94.4) | 82 (5.6) | <0.001 |
| Yes | 136 (78.2) | 38 (21.8) |  |
| Dyspnea<sup>a</sup> |  |  |  |
| No | 1401 (93) | 106 (7) | 0.087 |
| Yes | 114 (89.1) | 14 (10.9) |  |
| Initial Signs |  |  |  |
| Temperature °C<sup>b</sup> | 36.5 (36-37.2) | 37.5 (36.3-38) | <0.001 |
| ≤37.4<sup>c</sup> | 700 (93) | 53 (7) | <0.001 |
| 37,5-37,9 | 75 (83.3) | 15 (16.7) |  |
| ≥38 | 136 (78.2) | 38 (21.8) |  |
| SpO<sub>2</sub> %<sup>b</sup> | 95 (94-96) | 94 (93-95) | <0.001 |
| SpO<sub>2</sub> %<sup>a</sup> |  |  | <0.001 |
| ≥94% | 573 (92.4) | 47 (7.6) |  |
| ≤93% | 140 (76.9) | 42 (23.1) |  |
| PCR |  |  |  |
| Negative<sup>a</sup> | 474 (95.6) | 22 (4.4) | <0.001 |
| Positive | 353 (86.1) | 57 (13.9) |  |
| <sup>a</sup> Pearson's chi-squared test. <sup>b</sup> Mann–Whitney U test. <sup>c</sup> Chi Square for Linear Trend, <sup>d</sup> Fisher's exact test |  |  |  |

Table 3 shows Odds Ratio (OR) and its 95% confidence interval (95%CI) of the different symptoms for clinical deterioration unadjusted (bivariate analysis) and adjusted through a multivariate analysis in 5 steps model. Initially in the bivariate analysis the 5 symptoms resulted statistically significant: Fever OR 4.69 (95% 3.07-7.17, p<0.001), cough OR 3.16 (2.16-4.63, p<0.001), myalgias/arthralgias OR 2.7 (1.86-3.94, p<0.001), fatigue OR 2.56 (1.75-3.75, p<0.001), and headache OR 1.8 (1.23-2.61, p=0.002). Once they were adjusted among them and by demographic characteristics and comorbidity only four of them persisted statistically significance (Model 3): Fever OR 3 (1.89-4.77, p<0.001), 2), cough OR 2.34 (1.56-3.52, p<0.001), myalgias / arthralgias OR 1.69 (1.09-2.63, p=0.018), and fatigue OR 1.65 (1.08-2.53, p=0.019).

**Table 3.**
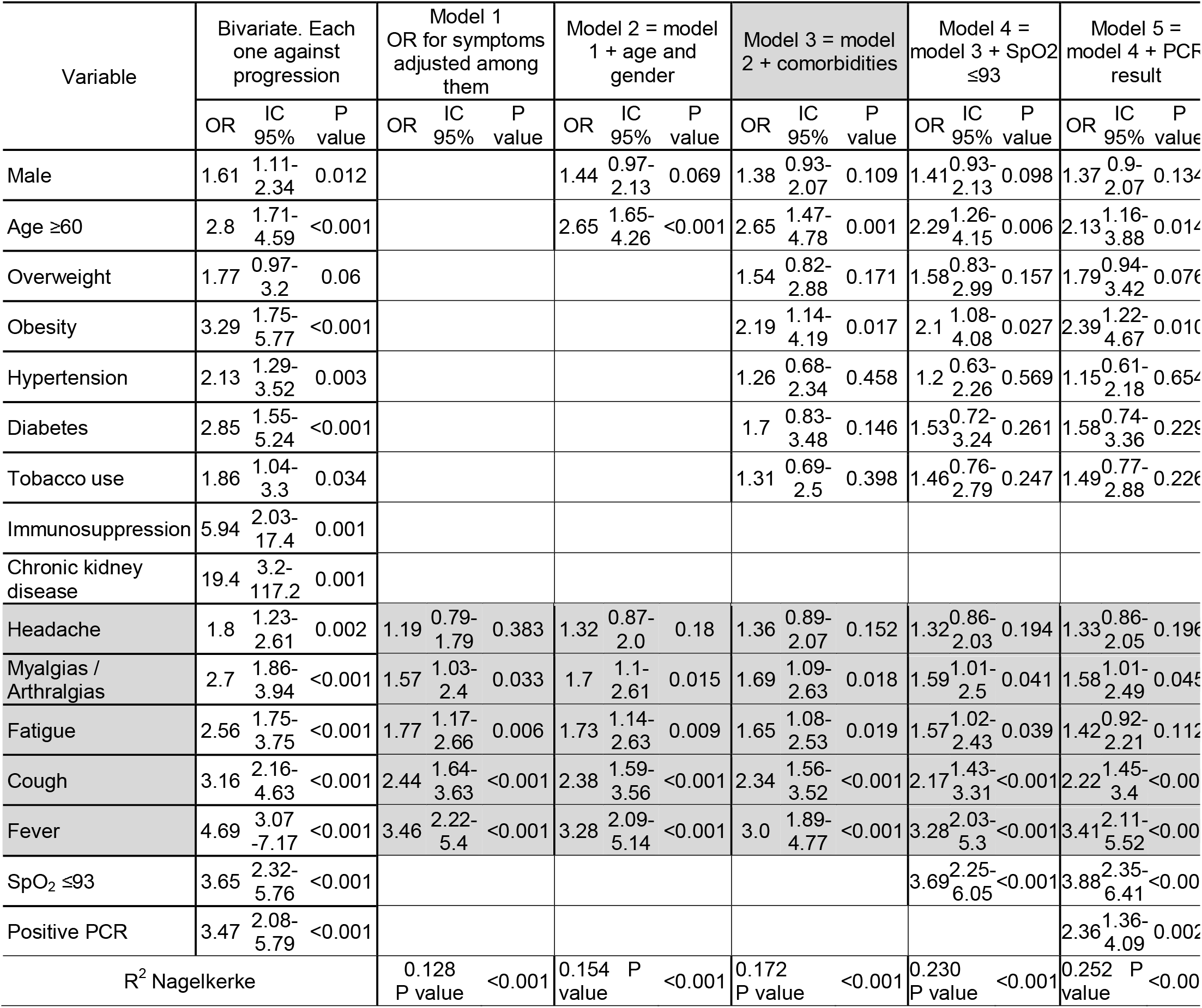
Multivariate analysis of Symptoms associated with disease progression in patients with COVID-19 suspicion.

Conjunctive consolidation analysis is shown in table 4 (process in supplementary table 1 and 2). Initially, because fever and cough had the highest OR were consolidated in the first step, then fatigue was added considering its higher gap, and a 3 groups score resulted, COVID-19 Progression According to Initial Symptom Index (C-19PAIS Index): 1) Fever with cough or fever with fatigue, with a probability of progression of 29% (31/106 patients), 2) Fever or cough or fatigue, or cough with fatigue, 10.7% (66/615 patients) and 3) No fever, no cough, no fatigue, 2% (23/914). Adding myalgias or arthralgias, only impacts on low-risk group, while presenting it the probability of progression increases from 2% to 7%, meanwhile the absence of it, decreases the probability to 1.3%.

**Table 4.** COVID-19 disease progression according to initial symptom (C-19PAIS Index)

| Progression risk | Progression n, (%) |
| --- | --- |
| High risk: Fever with cough // Fever with fatigue | 31/106 (29%) |
| Medium risk: Fever // Cough // Fatigue // Cough with fatigue | 66/615 (10.7%) |
| Low risk: No fever, no cough, no fatigued | 23/914 (2%) |

Adding myalgias or arthralgias, impacts on low-risk group: while presenting it the probability of progression increases from 2% to 7%, meanwhile the absence of it, decreases the probability to 1.3%.

Progression According to Initial Symptom Index (C-19PAIS Index) was contrasted against previously described prognostic risk factors in order to show its impact (Table 5): Among C-19PAIS Index, patients with high-risk suffer a great increment of progression when they are older (<60 years = 24% vs ≥60 years = 75%), and when they are obese (BMI <25 = 22%, 25-29.9 = 32%, ≥30 = 36%), diabetic (no = 28% vs yes = 40%), and tobacco use (no = 27% vs yes = 45%). Finally, when SpO2 measure is possible, lower values inclusive above 90%, represent an increased risk (SpO2 ≥94% = 29% vs >90 to ≤93% = 50%).

**Table 5.**
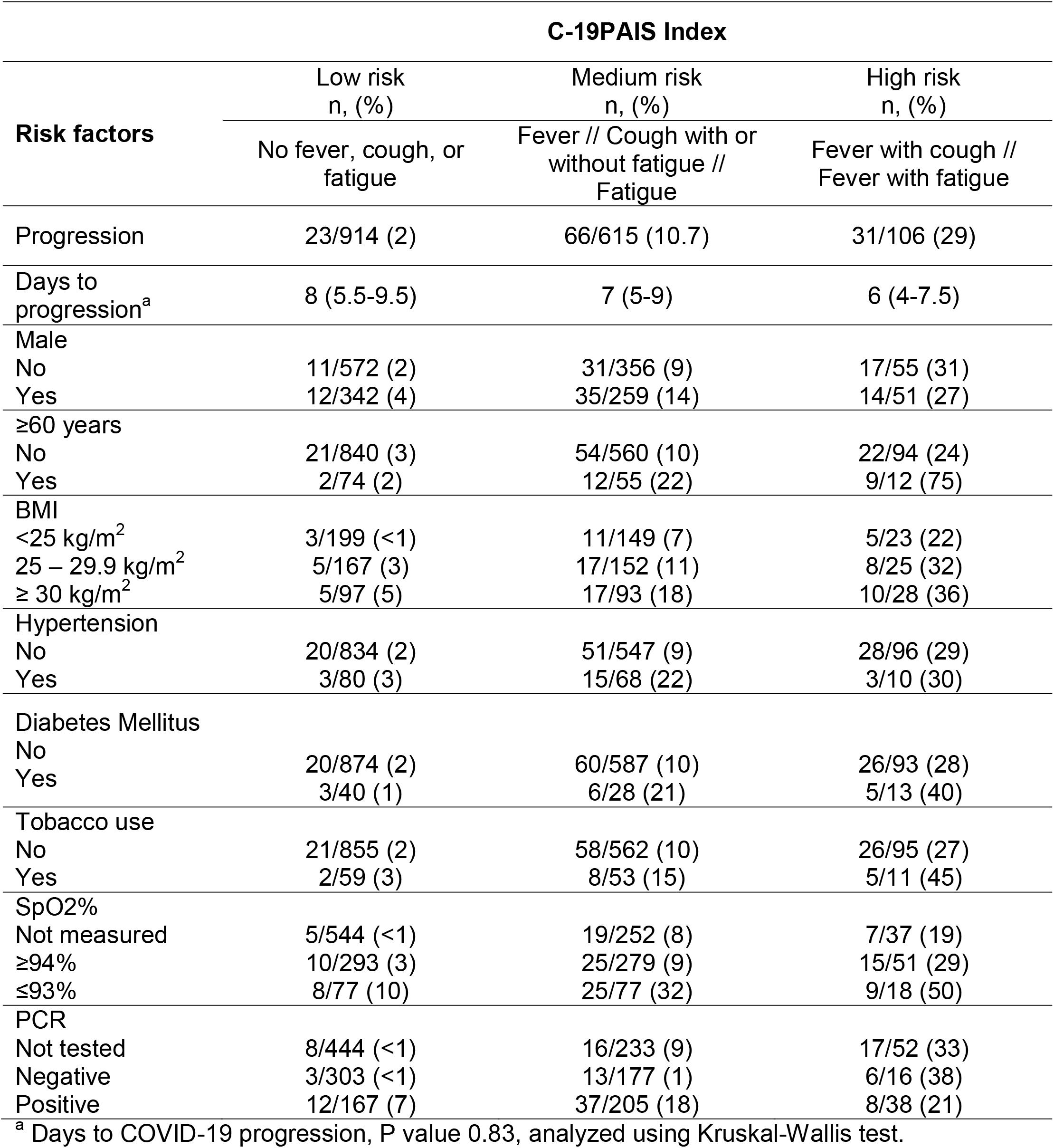
Progression at different stratums of C-19PAIS Index according to risk factors.

## Discussion

This study reported a progression to severe disease of 7.2% among patients with suspected or confirmed mild or moderate COVID-19 and asymptomatic person with recent contact with COVID-19 suspected or positive patients. Previous studies reported progression of 4.8%, 7.1%, and 19.2% (17-19), being up to 55% in moderate cases (20).

C-19PAIS Index can identify a group of patients with higher probability of progression to severe or critical disease by evaluating initial symptoms creating three different groups of patients: high-risk 29% (31/106 patients), medium-risk 10.7% (66/615 patients), and low-risk 2% (23/914).

Initial clinical presentation of disease with fever, cough, general malaise, and body aches, may be related to increase in inflammatory pathways. The immune response, after viral spike glycoprotein of SARS CoV-2 binds and merges through the cell membrane protein angiotensin-converting enzyme 2 (ACE2) of epithelial cells in the respiratory system(21), evoking innate immune system and activation of downstream cascades molecules and cytokines production, promoting cytokine storm, that is related to disease severity(22).

In other published studies, fever is less frequent in mild to moderate cases than severe or critical(23-25), and body temperature over 37.5ºC (13), is associated with higher risk of disease progression, as well as risk factor for severe, critical COVID-19, mechanical ventilation, and mortality (6, 13, 26-28). Higher levels of IL-1β and IL-6 were identified in patients with body temperature ≥38.0ºC (29), therefore fever as initial symptom may be related to earlier cytokine storm by means of increase in inflammatory pathways(30, 31).

Cough when evaluated as present or absent during course of disease, had demonstrated no difference in severity, being of the most frequently reported symptoms (20, 32, 33), however, as initial symptom was present in 48.2% of hospitalized patients that require supplementary oxygen (34).

Fatigue, myalgias and arthralgias, have been identified as symptoms related to COVID-19 progression (4), as well as other symptoms not evaluated in this study, for instance chills (11) with and increased risk for progression to severe disease, or sputum, which represented a risk factor for patients with mild or moderate disease to high flow oxygen therapy and mechanical ventilation (15). Headache, despite being more frequent in the progression group, is not a risk factor for clinical aggravation, as stablished previously (4).

As mentioned before, systemic symptoms and feverish patients predict clinical deterioration (25), therefore, it is not a surprise, that clinical initial symptoms resulted useful to detect those patients who would present disease progression.

Demographic and clinical characteristics of high impact for severe disease like male, older age, obesity, and comorbidities had been reported as risk factors (3, 6, 7, 26, 35-37). In our study, being male, as shown in previous studies resulted associated with increased risk progression in bivariate analysis, however, when adjusted for age and symptoms, had no impact; it could be the result of higher age among men or that clinical symptoms identify in a better way those men that will progress.

It is also important to highlight an increased risk at the interior of high-risk strata of C-19PAIS Index in older patients (75% vs 24%) and some comorbidities like obesity (36% vs 22%), diabetes mellitus (40% vs 28%), hypertension (sum of medium and high-risk group, 23% vs 12.2%), and tobacco use (45% vs 27%). Blood tests, like white blood cell count, lymphocyte count, C reactive protein, D dimer, ferritin, and IL-6 (6, 36, 38), a positive SARS CoV2 PCR (9, 36) and radiology manifestation (7, 13, 39-41) are some paraclinical studies which may predict aggravation and illness severity, however, all of them require people going out for medical service while in asymptomatic people with positive exposition, and symptomatic with mild or moderate disease this strategy of follow up trough symptoms at home results attractive and efficient. Patients with C-19PAIS Index in high-risk should be evaluated in a close manner through symptomatology -twice a day-, and in cases of being older, or with obesity, diabetes, hypertension, or tobacco, paraclinical tests like SpO2% by pulsioximeter may be added. Those in middle-risk group should be monitored daily, and low-risk group, due to its low probability of progression, may be monitored every two days. In all risk-group patients, must be instructed about alarm symptoms and referred immediately to hospital evaluation when these were present.

This project has several strengths, but it should be noted that the main one is that it reflects a real option in homecare for cases in which hospital or office-level care is exceeding or involves a risk. Of course, it can only be considered in diseases where there are low-risk groups and a type of information that can be collected through a telephone interview with a patient or caregiver capable of responding.

This study has limitations. Clinical interview was realized by many doctors with different training (general practitioners, internists, and otorhinolaryngologists), although, all of them received a standardized training to participate in the program.

C-19PAIS Index may help discern those with low from high risk of progression, identifying those who require more intensive monitoring and extension tests, anticipate, and prevent adverse outcomes.

## Data Availability

All data produced in the present study are available upon reasonable request to the authors

