## Supplementary table 1 and 2 for "COVID-19 disease progression according to initial symptoms. A telemedicine cohort study"

**Supporting information**

S1 Table. Conjunctive consolidation step 1; consolidation of fever and cough: progression proportions of COVID-19 suspected or confirmed patients.

| **Supplementary Table 1 Conjunctive consolidation step 1; consolidation of fever and cough: COVID-19 disease progression according to initial symptom (C-19PAIS Index)** | | | |
| --- | --- | --- | --- |
| Fever | Cough | |  |
|  | Yes (%) | No (%) | Total (%) |
| Yes (%) | 19/61 (31) | 19/113 (17) | 38/174 (21.8) |
| No (%) | 35/304 (11.5) | 47/1157 (4) | 82/1461 (5) |
|  | 54/365 (14.8) | 66/1270 (5) |  |

Four clusters are formed, a) fever with cough, b) fever without cough, c) cough without fever,

d) no fever and no cough.

S2 Table. Conjunctive consolidation step 2; consolidation of fever and cough, with fatigue

| **Supplementary table 2. Conjunctive consolidation step 2; consolidation of fever and cough, with fatigue: COVID-19 disease progression according to initial symptom (C-19PAIS Index)** | | | |
| --- | --- | --- | --- |
| Fever and cough clusters | Fatigue | |  |
|  | Yes | No | Total |
| Fever with cough | 6/24 (25) | 13/37 (35) | *19/61 (31)* |
| Fever without cough | 12/45 (26) | *7/68 (10)* | *19/113 (17)* |
| Cough without fever | *12/108 (11)* | *23/196 (12)* | 35/304 (12) |
| No fever and no cough | *24/243 (9)* | 23/914 (2) | 47/1154 (4) |
|  | **Composites zones** | |  |
|  | 31/106 (29%) |  | High risk |
|  |  |  | Medium risk |
|  | *66/615 (10.7%)* | |  |
|  |  | 23/914 (2%) | Low risk |

High risk: fever with cough or fever with fatigue. Medium risk: fever alone or cough alone or fatigue alone, or cough with fatigue. Low risk: no fever, no cough, and no fatigue.
